# Personalized and dynamic antibiograms-an exploration in seven infectious syndromes

**DOI:** 10.1101/2021.01.22.21249954

**Authors:** Michelle J. Iandiorio, Jane C. Fazio, Prakasha Kempaiah, Ravi Darvasula, Marc H. V. van Regenmortel, Ariel L. Rivas

## Abstract

To prevent antimicrobial resistance and inform better, antibiograms should distinguish different biomedical situations. It is also desirable that new antibiograms provide *in vivo*, temporal, and patient-specific immunological information. Here, the informative ability of a pattern recognition-based method was explored with data collected from patients that experienced seven infectious syndromes (pneumonia, endocarditis, tuberculosis, syphilis, as well as skin and soft tissue, intra-abdominal, and/or urinary tract infections associated with meningitis). Interactions among seven dimensions (7D) were investigated: (i) space, (ii) time, (iii) temporal data directionality, (iv) immunological multicellularity, (v) antibiotics, (vi) immunomodulation, and (vii) personalized data. Omissions and ambiguity (confounding different biological situations) occurred when static metrics were used in isolation, such as leukocyte percentages. In contrast, hidden information was uncovered when complexity and dynamics were assessed. The 7D approach grouped together observations that displayed similar immune profiles and identified antibiotics that modulated specific leukocytes. For instance, in tuberculosis, blood monocytes were modulated by isoniazid-related antimicrobials. In spite of the diverse syndromes analyzed, this proof-of-concept discriminated. It is suggested that the simultaneously exploration of numerous dimensions associated with complexity may be biologically interpretable, prevent ambiguity, promote research, expand machine learning-oriented methods, and support personalized medicine.

## INTRODUCTION

New methods have been recently requested by physicians. Such calls respond to the ‘reproducibility crisis’ of clinical research, characterized by up to 90% of irreproducible studies.^1^ Numerous factors seem to cause this problem, which are not limited to statistical errors.^2^ They also include reductionist practices, e.g. (i) the ‘streetlight’ effect (measuring what is easily measured, but not biomedically relevant), (ii) dichotomous and static methods applied to biomedical conditions that may be dynamic and polychotomous; and (iii) research designs that ignore co-morbidities and interactions.^1-7^

Because the unit of interest, in clinical medicine, is one patient (not a plurality, as in population medicine), neither averages nor linear models apply and, consequently, it cannot to be established whether one specific patient will benefit from a specific intervention.^2, 8, 9^ Personalized medicine also requires novel cognitive approaches. The current emphasis (promoted by ‘evidence-based’ medical practices) expects clinicians to reason *from the general (population average) to the particular* (*patient)*. However, the history of each patient (including multi-morbidities and polypharmacy) also needs be considered.^10, 11^

To address these issues, non-reductionist analyses of biomedical complexity and dynamics are needed. Because two patients receiving the same diagnosis may differ in outcomes, prognosis should also be explored. ^2, 4, 9, 12, 13^ Future methods should not utilize a single type of data but integrate theory with data that includes spatial-temporal dimensions.^14-17^ In addition, ‘one person trials’ may be considered, which may investigate *immunological multicellularity* –a critical component in responses against infections.^18-20^

Antimicrobial resistance is one example of the medical problems non-reductionist approaches should address. New strategies could go beyond the classic focus on antibiotic-pathogen relationships and also interrogate the immune response of the host.^21,22^

Research on immunomodulation has shown, for example, that norfloxacin promotes neutrophils, while linezolid suppresses phagocytosis, both *in vivo* and *in vitro*.^23, 24^ Similar studies have shown that macrolides suppress bacterial infection of the lungs and reduce inflammation.^25^ Long-acting macrolide antibiotics, such as azithromycin, can protect even if given intermittently.^26^ To prevent multi-drug antimicrobial resistance, combinations of antibiotics and antibodies have been proposed.^27^

Studies that investigate immunological multicellularity and/or immunological and antibiotic interactions matter clinically because, in many infections, empirical antibiotic treatments are prescribed when the identity of the pathogen is unknown. For instance, in pneumonia, pathogens may be isolated in only one third of the cases.^28^ Thus, earlier and *in vivo* assessments are needed to evaluate the efficacy of empirical treatments.

Investigations on antibiotic-immunological interactions could also shed light on the pathogenesis of major diseases and, consequently, influence vaccine development. For instance, very little is known on the pathogenesis of tuberculosis –a disease that requires better vaccines because current ones have only a 30% efficacy.^29, 30^ In particular, immunological-antibiotic interactions should elucidate whether antibiotics foster or inhibit the functions of immune cells.^31, 32^

Since 1947, antibiotics have been investigated with *in vitro* tests (antibiograms or antibiotic sensitivity). Such tests have several limitations, including absence of dynamic perspectives, which only *in vivo* studies can provide.^33-35^ Because they lack personalized information, these tests inherently support the ‘one dose fits all’ paradigm.’^36^

While the problems associated with reductionism have been abundantly documented over two decades^37-39^, very few solutions have been proposed. Here an approach meant to ameliorate reductionism-related problems is advanced.

To materialize this pursuit, novel tests used in infectious diseases should demonstrate that they do not generate ambiguous results. Ambiguity refers to lack of decision-making not due to lack of data but because the data do not discriminate.^40^ Health *trajectory* (also known as *temporal data directionality*) is one strategy that may prevent ambiguity.^41^ Trajectory differs from classic measurements of time: by using arrows that connect pairs of consecutive observations, the *directionality* of temporal data is unmasked, providing a new level of information. Health trajectory is a person-centered metric that describes the ‘flight path of an object’, informing on the direction and/or speed of a health change. While trajectory has been studied with aggregate data, the complex dynamics of health trajectories have not been explored at personalized bases.^42, 43^ While trajectory has been investigated in infections, earlier studies did not assess antibiotics.^40, 44^

Data collected from patients affected by seven infectious syndromes were explored. They included endocarditis, pneumonia, tuberculosis, syphilis, as well as intra-abdominal, skin and soft tissue, and/or urinary tract infections associated with meningitis. The selection of syndromes was based on their prevalence, in-hospital mortality, and/or recent trends. For example, in the US, infective endocarditis results in prolonged hospitalization and is associated with 20% in-hospital mortality.^45^ Worldwide, pneumonia causes 2-3 million annual deaths and, in the US, approximately 100,000 in-hospital annual deaths.^46,47^ Intra-abdominal infections may result in up to 36 % mortality.^48^ While rarely fatal, the prevalence of skin and soft tissue infections is rapidly increasing in the US.^49^

Seven dimensions were explored: (i) three-dimensional (3D) space, (ii) time, (iii) trajectory, (iv) multicellularity, (v) antibiotics, (vi) immunological-antibiotic interactions, and (vii) patient-specific information. The goal of this study was to elucidate (i) whether ambiguity was a rare phenomenon, and (ii) if not, whether a non-reductionist approach could prevent ambiguity, discriminate, and/or inform.

## MATERIALS AND METHODS

### Data

Seventy longitudinal observations collected from nine adults diagnosed with infective endocarditis, pneumonia, tuberculosis, syphilis, intra-abdominal, skin and soft tissue, and/or urinary tract infections associated with meningitis were analyzed. Day 0 indicated the first consultation or the date of hospitalization.

This study was conducted as described in Protocol #13-463, which was approved by the Institutional Research Protection Office committee of the Health Sciences Center, University of New Mexico on June 23, 2016 (protocol titled ‘Small Dataset-Based Discrimination of Infectious Disease Pattern’, Dr. M Iandiorio, PI). This protocol protects the identity of the patients investigated. The data were anonymized by a double-blind process in which the treating clinician does not participate in the analysis of the data and the data analyst does not receive any information that could identify the patients. Because this protocol allows the retrieval and analysis of clinical data from historical clinical records without a previous review, no waiver was requested and no decision from the competent authority was required in advance for any one study. The identification numbers reported in this study were created by the analyst in order to perform the analysis –they do not / cannot identify any person.

### Assessment of biomedical validity

Construct, internal, and external validity were investigated. Statistical validity was not determined because such a research goal is only justified after the other types of validity are shown to be defensible. Because external validity depends on or follows construct and internal validities, demonstration of external validity was emphasized.^50^

This study was designed to determine whether the data patterns, if any, were robust to both patient and syndrome variability. To that end, a dataset was constructed to include: (i) at least five infectious syndromes and nine patients; (ii) at least two infectious syndromes would be explored with data from two patients, (iii) at least one co-morbidity would be investigated; (iv) at least one patient was not treated with antibiotics; and (v) in at least one patient, longitudinal observations would not start with antibiotic treatments so pre-/post-treatment data could be investigated. External validity was regarded to be plausible when at least one multicellular interaction (e.g., the lymphocyte % over the monocyte % or L/M ratio) distinguished non-overlapping data subsets, patients, trajectories, temporal phases and/or infectious syndromes.

### Analyses

Distinct data patterns were identified as described elsewhere.^4, 13, 40, 44^ Partitioning into data groups that, internally, exhibited similar immune profiles, was conducted using a proprietary algorithm (US patent 10,429,389; 2019). Two- and three-dimensional (2D and 3D) plots were created with a commercial package (Minitab Inc., State College, PA, USA).

## RESULTS

### Detection and prevention of ambiguity

Analyses of individual cell types were ambiguous. For example, a patient diagnosed with endocarditis exhibited similar leukocyte percentages even when samples were collected 29 days apart (Figs. 1 A, B). Because leukocyte percentages do not explore interactions, further analyses measured dimensionless indicators (DIs) designed to capture complexity. When time was investigated, DIs revealed numerous changes in trajectory, which differentiated two subsets that exhibited: (i) ‘left-to-right’, and (ii) ‘right-to-left’ flows, respectively (Figs. 1 C, D). Trajectory-based assessments showed non-overlapping data intervals of interpretable variables: when the lymphocyte percentage was divided over the monocyte percentage (the L/M ratio), two subsets were distinguished (Fig. 1 E). When spatial patterns and multicellular interactions were analyzed, the 7D method differentiated (i) two data subsets, as well as (ii) three data points that, previously, could not be separated (Figs. 1 F, G). Complex multicellular indices –the L/M ratio and an indicator of greater complexity (e.g., the [L/M]/[N/L] ratio)− discriminated more or better than leukocyte percentages (Figs. 1 B, G).

**Figure 1.**
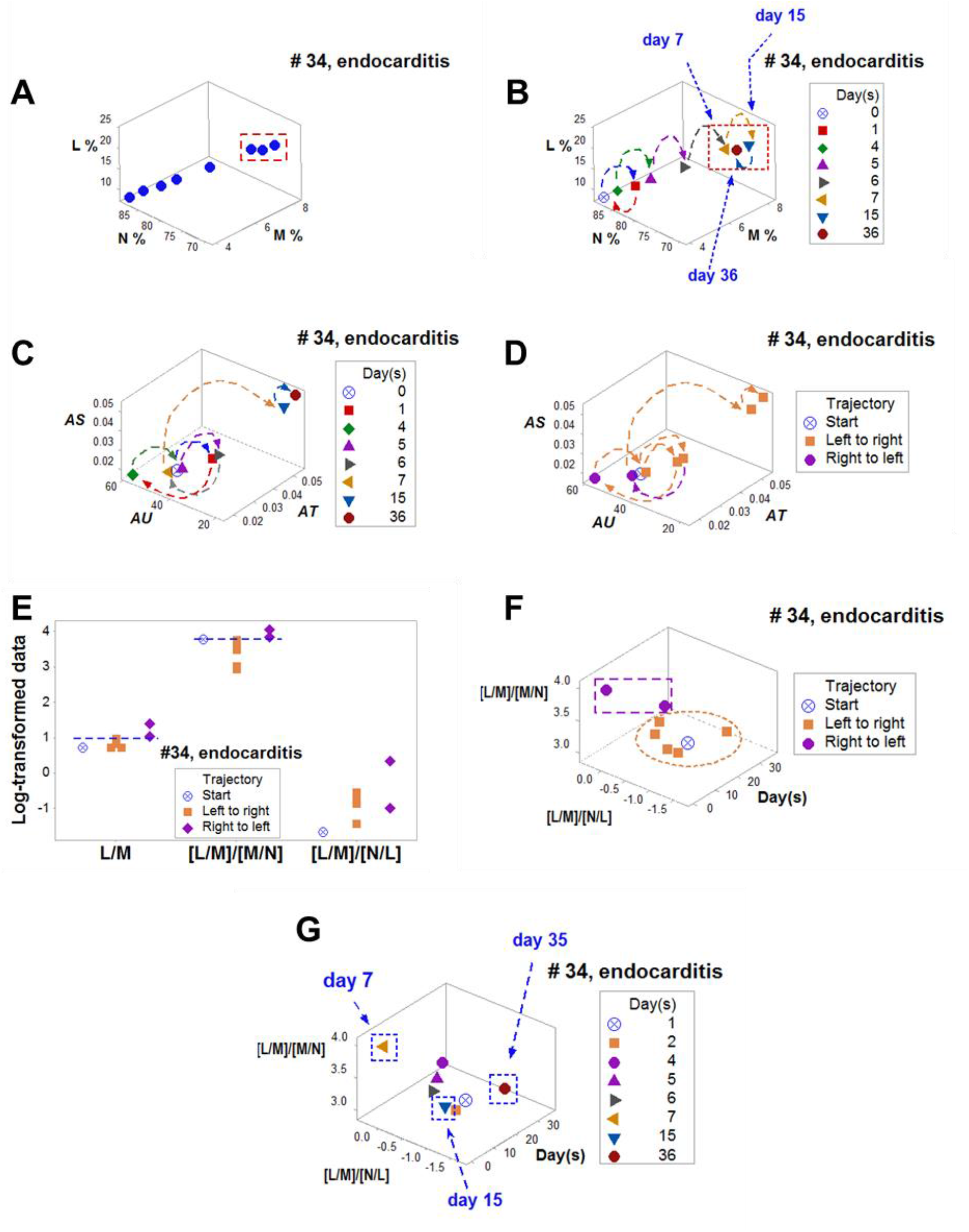
Detection and prevention of ambiguity in endocarditis. Revealing ambiguity, three numerically similar observations were found at different time points (**A, B**). Over time, dimensionless indicators differentiated two data directionalities (trajectories): from ‘left-to-right’ and from ‘right-to-left’ (**C, D**). Trajectory distinguished two data subsets, which displayed non-overlapping intervals of L/M and [L/M]/[M/N] ratios both when time was not and was considered (**E, F**). When space, time, and multicellularity were analyzed, the three data points previously regarded as ambiguous were clearly differentiated (**G**).

Data collected from another patient diagnosed with endocarditis and treated with antibiotics highlighted the dynamics of the immunological-antibiotic interactions: differentiating two temporal phases, vancomycin and cefazolin both promoted and inhibited immune responses (SI Figs. 1 A-H). The discriminant ability of the L/M ratio was, again, documented: non-overlapping data intervals separated an earlier from a later data subset, even when the M percentage exhibited overlapping intervals (SI Figs. 1F, H).

### Short-term ambiguity and real-time antibiotic monitoring

Skin and soft tissue infections (SSTI) also revealed ambiguity when investigated with isolated variables (Figs. 2 A-D). When the neutrophil/monocyte (N/M) and the lymphocyte/monocyte (L/M) ratios were explored, data points collected at the third and fourth day were assigned to separate clusters (Fig. 2 E). In contrast, data points recorded nine days apart were clustered together (Fig. 2 E).

**Figure 2.**
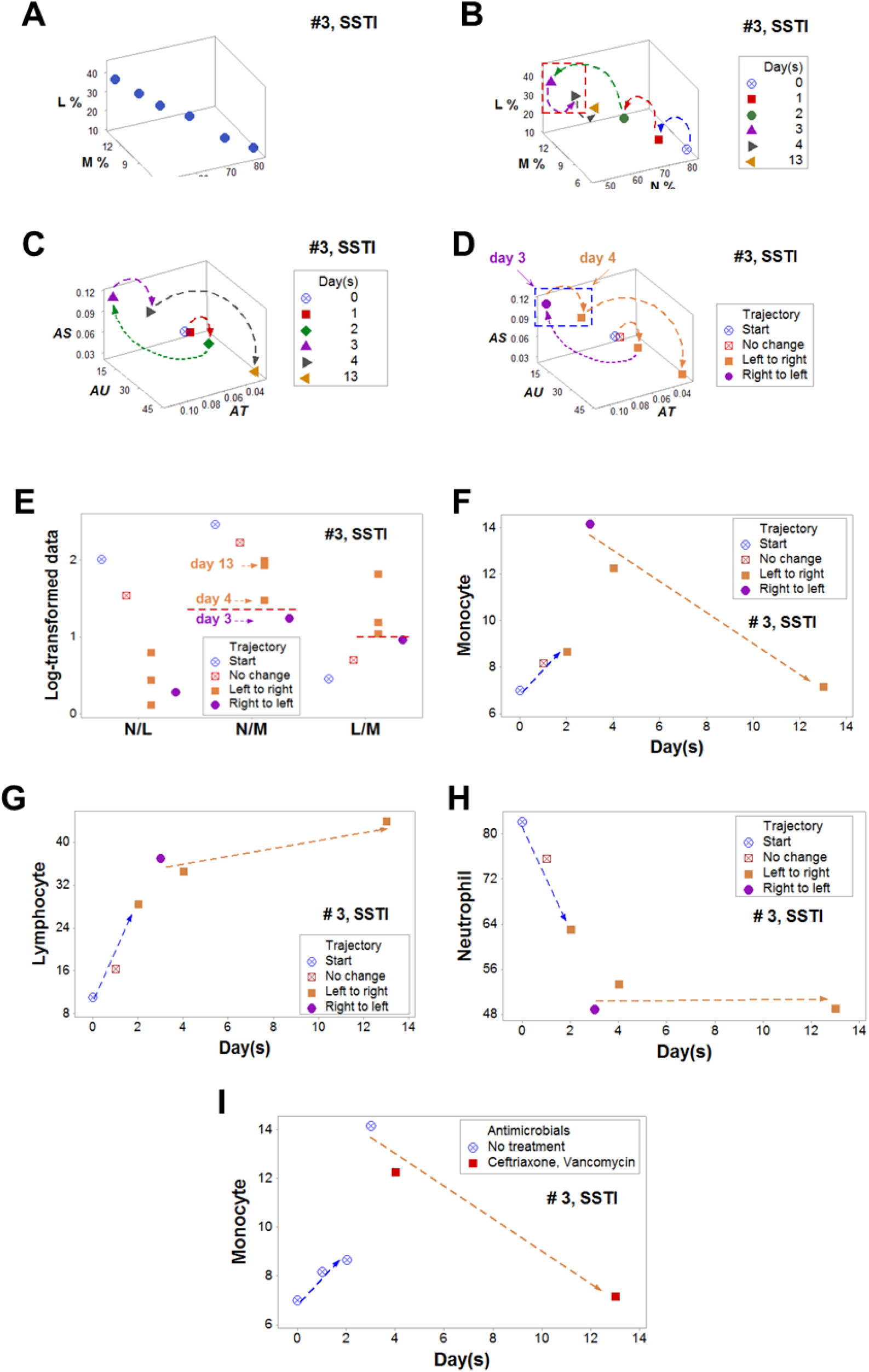
Detection and prevention of ambiguity and antibiotic modulation of monocytes. Ambiguity was also found in a case of skin/soft tissue infections (**A, B**). The analysis of immunological complexity and trajectory identified two data subsets, including observations recorded one day apart (which were assigned to different subsets) and observations collected 9 days apart (which were clustered together, **C-E**). When time, trajectory, and specific leukocyte cell types were considered, two subsets were distinguished which did not include or included information on antibiotic therapy (the earlier and later subsets, respectively, **F-I**). Because only the monocyte percentage data displayed subsets orthogonal to one another and showing opposite directionalities, findings supported the hypothesis that ceftriaxone and vancomycin, together, modulated monocytes (**F-I**).

When trajectory was considered, the M percentage revealed two data subsets orthogonal to one another, which exhibited different directionality (Fig. 2F). While the L and N percentages also displayed two subsets, they were not perpendicular to one another (Figs. 2 G, H). Because only the M percentage changed directionality after antibiotic treatment, it was concluded that ceftriaxone and vancomycin immunomodulated monocytes (Fig. 2 I).

### Pattern reproducibility

A second SSTI case demonstrated that ambiguity may occur even when temporal data directionality is measured (SI, Figs. 2 A-H). While uni-dimensional (1D) analyses of percentages could be ambiguous (SI, Figs. 2 E, F), 3D analyses of complex interactions discriminated (SI Fig. 2G).

The N/M ratio was demonstrated to be a well conserved function: it informed in two SSTI patients (Figs. 2 and SI 2). Within one day, the complex [N/M]/[L/M] ratio indicated recovery after nafcillin was administered (SI, Fig. 2H).

Ambiguity was also found in pneumonia: two data points were ambiguous when the neutrophil and monocyte percentages were evaluated (Fig. 3 A, B). In contrast, the analysis of complexity, dynamics, and trajectory identified two non-overlapping data subsets (Figs. 3 C-F). The pattern recognition/based method discriminated earlier (at day 9, when the N/L ratio was used [Fig. 3 G]) than when the N% was measured (Fig. 3 A) as opposed to day 18 and indicated that recovery was not initiated by any one antibiotic (Fig. 3 H).

**Figure 3.**
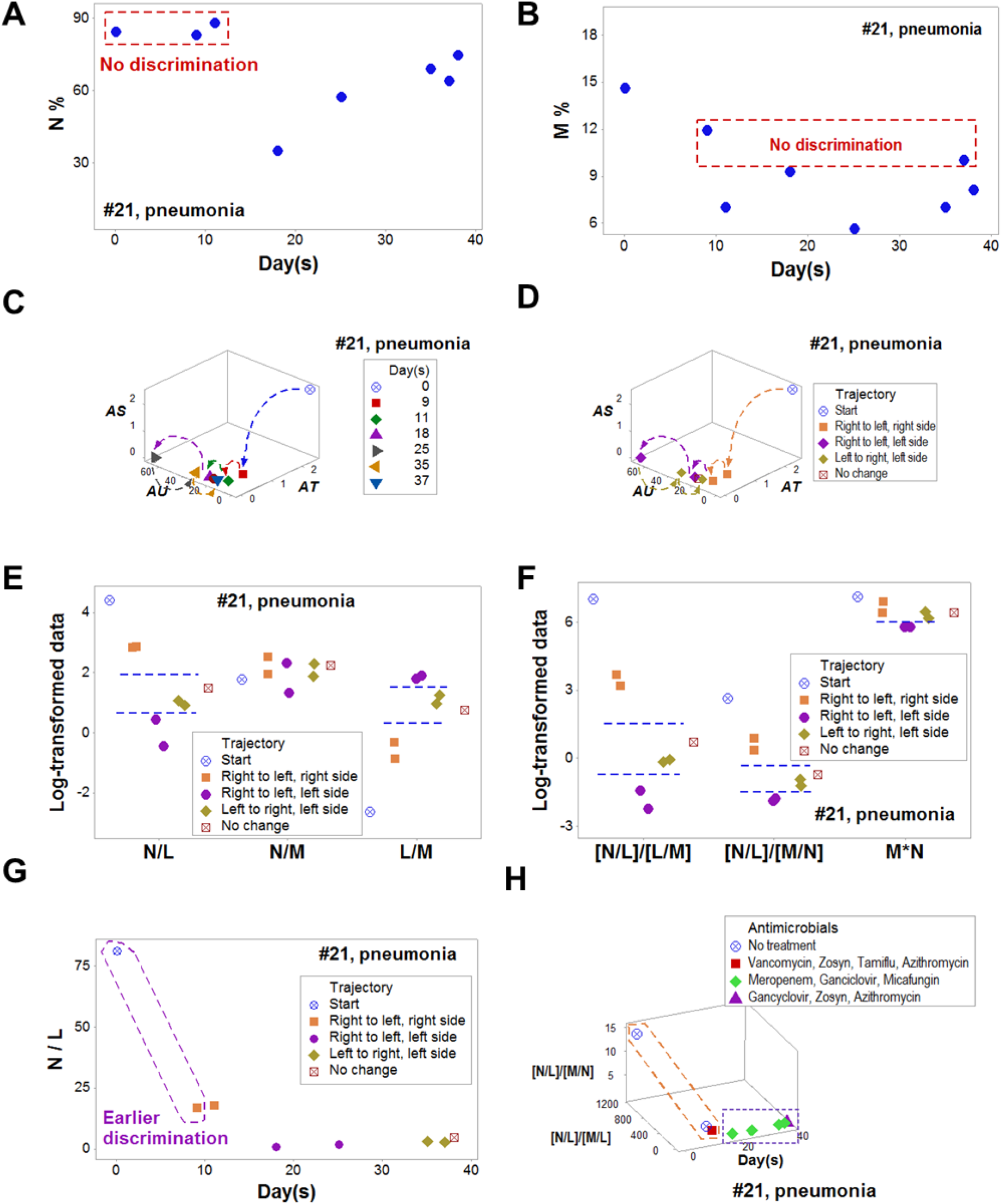
Earlier discrimination in pneumonia. Ambiguity was documented twice when leukocyte percentages were analyzed over time (**A, B**). The analysis of complexity, time, and trajectory revealed two non-overlapping data subsets (**C-F**) which: (i) discriminated earlier (at day 9, when the N/L ratio was used [**G**], as opposed to day 18 when the N% was measured, A), and (ii) indicated that recovery was not initiated by any one antibiotic (**H**).

Multidimensional analysis discriminated two subsets of intra-abdominal infections (Figs. 4 A-E). In contrast, the analysis of individual cell types failed to distinguish data subsets (Figs. 4 F-H). Ceftriaxone and metronidazole appeared to modulate a complex function characterized by the [L/M]/[N/L] ratio (Fig. 4 E).

**Figure 4.**
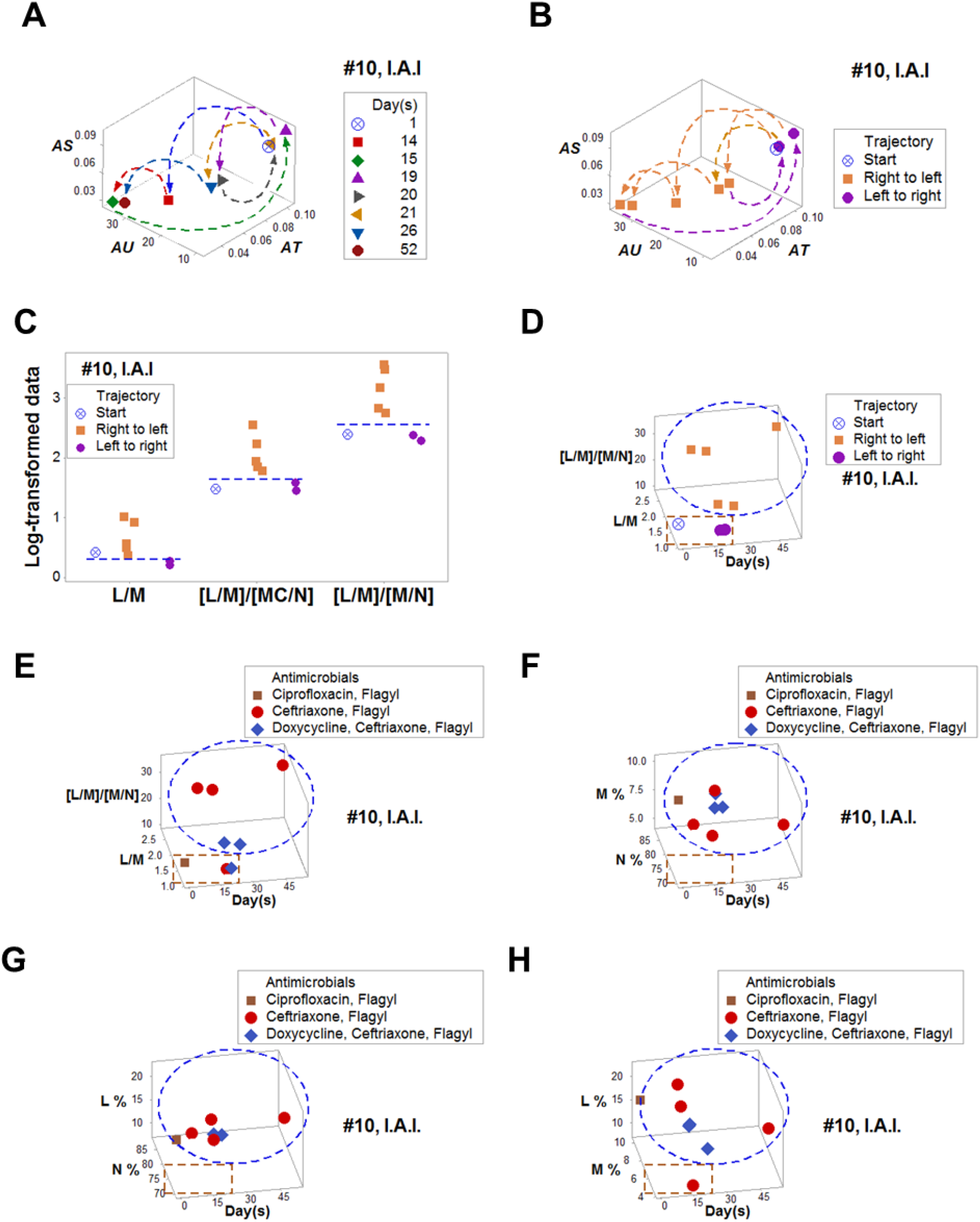
Multicellular modulation in an intra-abdominal infection. The analysis that included multicellular complexity, trajectory, time, and antibiotics in space discriminated two data subsets (**A-E**). In contrast, the analysis of individual cell types did not discriminate (**F-H**). Ceftriaxone and metronidazole modulated not a single cell type but a complex function characterized by the [L/M]/[N/L] ratio (**E**).

### Redundancy and immune-modulation in tuberculosis

Discrimination did not depend on any one dimensionless indicator: three different group of DIs (the L/M, [L/N]/[M/N], and [L/M]/[M/L] ratios) differentiated two 3D data subsets (Figs. 5A-D). In contrast, leukocyte percentages did not discriminate even when tested in pairs (Figs. 5 E-G). Yet, when antibiotics were measured, both 3D and 2D analyses revealed that monocytes were modulated by the isoniazid-related antibiotics (Figs. 5 H, I).

**Figure 5.**
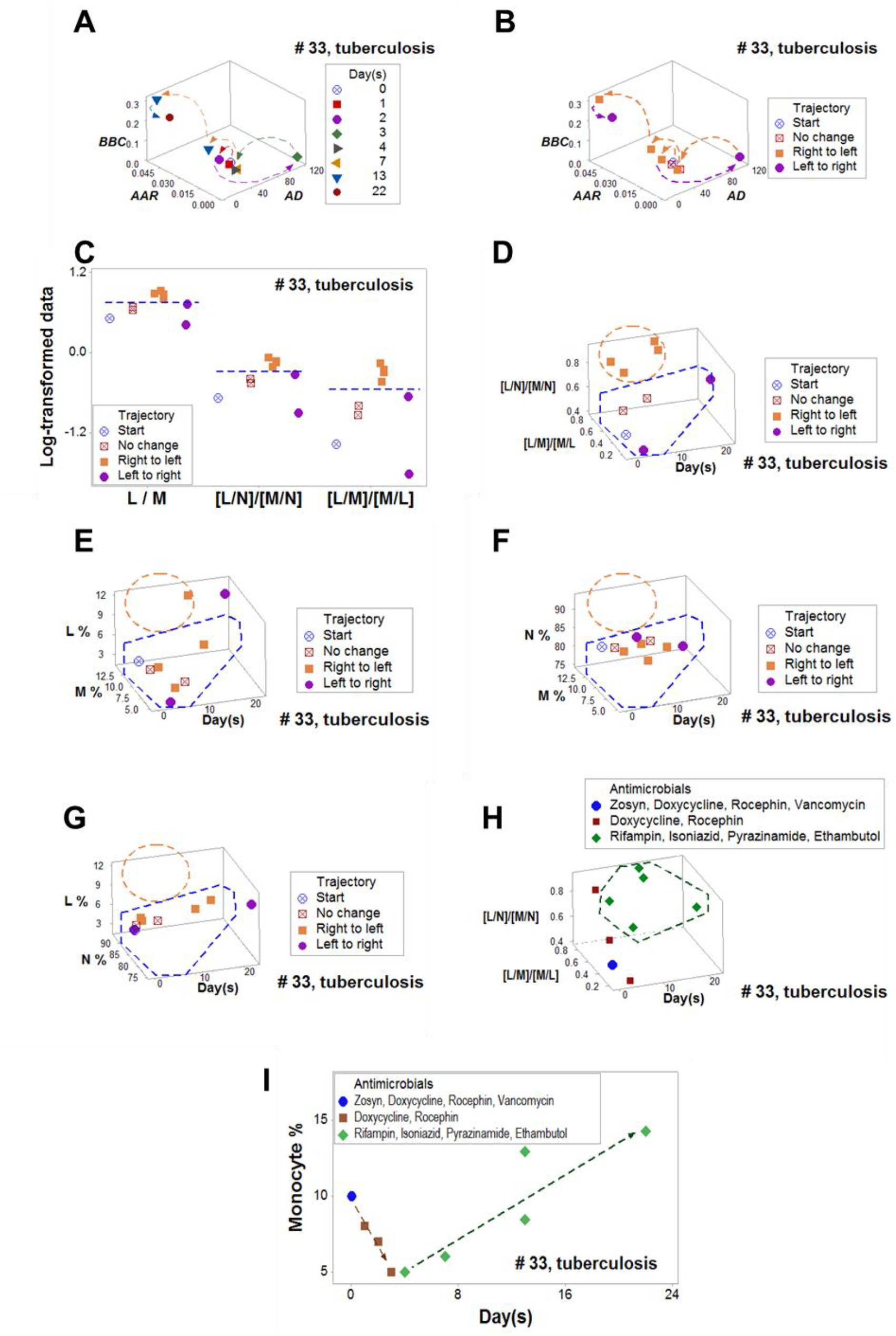
Monocyte modulation in tuberculosis. While a multidimensional analysis distinguished two subsets that revealed non-overlapping values (**A-D**), no pair of leukocytes, when measured as percentages, identified such subsets (**E-G**). An additional subset was identified when antibiotics were measured, which was associated with isoniazid-related therapy (**H**). When the trajectory of specific cell types and antibiotics were tested, isoniazid-related therapy seemed to modulate monocytes (**I**).

Ambiguity was also observed in neurosyphilis (boxes, SI, Figs. 3 A, B). When complexity and trajectory were measured in space/time, two data subsets were distinguished, which differed in both L/M and [L/M]/[M/L] ratio values (SI, Figs. 3 C-E). When trajectory was assessed, these metrics distinguished three subsets (SI, Fig. 3 F). When antibiotics were considered, the 3 subsets were reconfigured, indicating that clindamycin, aztreonam and trimethoprim/sulfamethoxazole modulated a complex function characterized by the [L/M]/[M/L] ratio (SI, Fig. 3 G).

### Feedback patterns in co-morbidities

Ambiguity was associated with co-morbidities (Fig. 6A). When space, time, immunological multicellularity, and trajectory were investigated, three temporal phases were observed, which revealed circular (feedback-like) data patterns (Figs. 6 B-E). When antibiotics were also investigated, amphotericin B was the only antifungal associated with all temporal phases (Fig. 6 F). When leukocytes were explored over time, a ‘V’ shape pattern supported the notion that lymphocytes were modulated by amphotericin B (Figs. 6 G-I).

**Figure 6.**
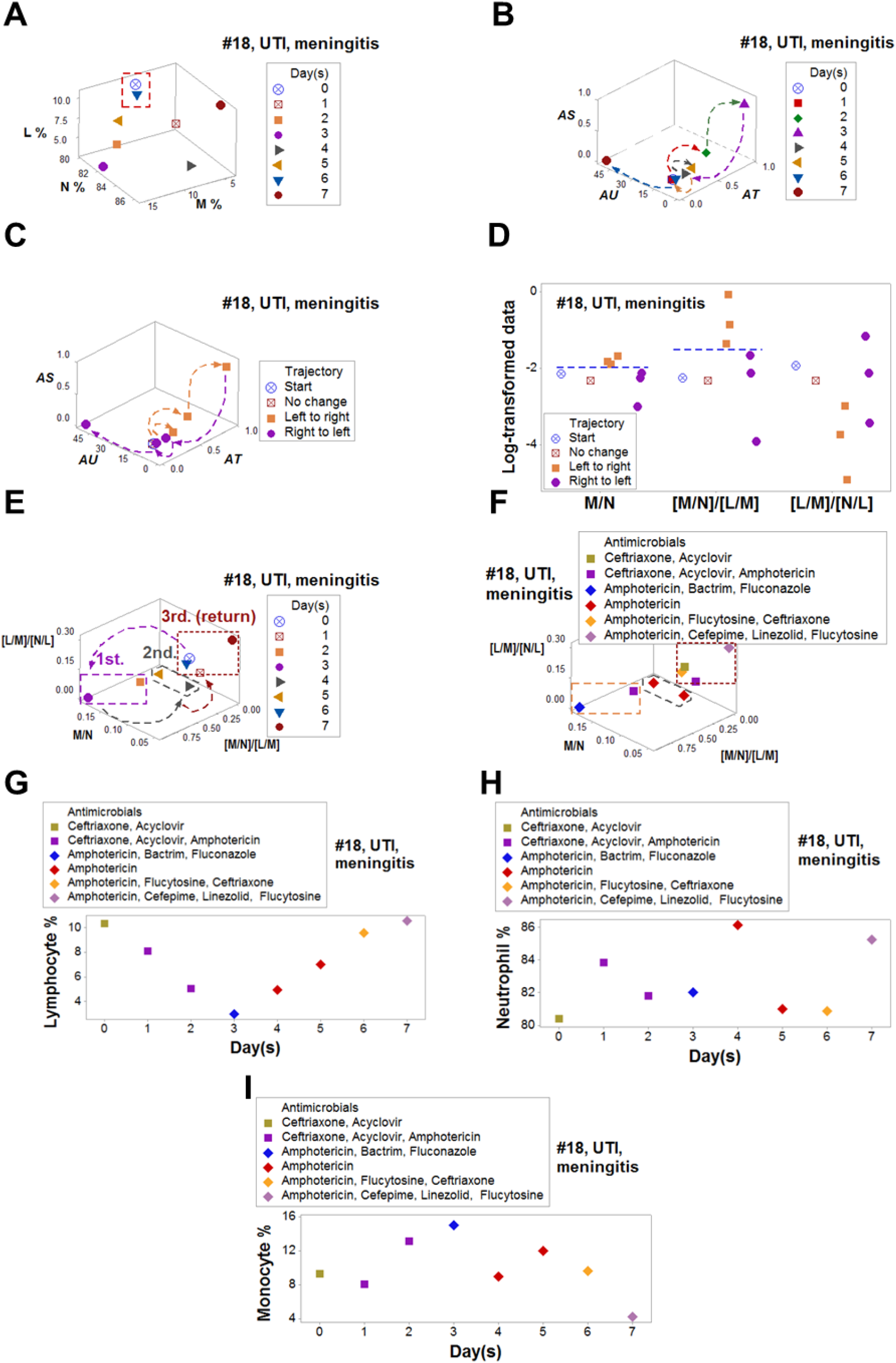
Lymphocyte modulation in multi-morbidity. Ambiguity was expressed as two temporal observations, collected six days apart, that expressed similar values of lymphocyte (L), neutrophil (N) and monocyte (M) percentages (**A)**. However, the combination of space, immunological complexity, trajectory, time and antibiotics distinguished three non-overlapping data subsets that expressed feedback (a circular pattern, **B-E**). Amphotericin characterized all data points included in the last subset (**F**), which modulated lymphocytes but no phagocytes (**G-I**).

## DISCUSSION

Nine out of 9 assessments revealed various expressions of ambiguity. They were prevented and more or novel information was extracted when seven dimensions were investigated. Findings are discussed along methodological and clinical perspectives.

### Methodological considerations

Ambiguity was observed in all patients. Given its ubiquity, it is suggested that ambiguity may contribute to the reported ‘reproducibility crisis’ of published research.^1^

Ambiguity is related to synergism and pleiotropy: the same structure (e.g., a cytokine or a cell) can perform many (and even opposite) functions, at different times. Monocytes/ macrophages illustrate how ambiguity occurs: they promote neutrophil activity at the beginning of the immune response and, a few days later –when the early inflammatory stage ends–, they destroy neutrophils.^51^ Therefore, a ‘10% monocyte’ value can be ambiguous because it can occur in different biomedical conditions or times, as shown in Fig. 3B.

Because infection-related data are, at least, four-dimensional, ambiguity is mathematically intractable.^13, 52, 53^ Any biomedical variable may have a very large number of combinations (if not infinity) when all dimensions are considered, i.e., for any value of ‘width’, there are many possible values of ‘height’, ‘depth’, and ‘time.’ Yet, ambiguity may disappear when context is added.^53^ Here contexts were explored considering (i) temporal interactions (the dynamics of immunological multicellularity with and without antibiotics) and (ii) other dimensions.

The information provided by the non-reductionist approach may result from the complexity and dynamics of multi-scalar and multicellular interactions. When changes occur at small temporal scales –e.g., molecular processes that take place at a nanosecond scale–, any study based on a larger scale (e.g., days) will miss such changes.^54^ These errors are prevented when temporal data directionality is measured: arrows pointing at different directions indicate changes. In addition, the simultaneous analysis of up to seven dimensions can generate richer information on multi-level, inter-dependent biological interactionss.^55^ While the analysis of complexity is not always informative, finding demonstrated that dynamics (longitudinal data on leukocytes and antibiotics) both informed and explained.

The 7D approach is compatible with classic statistics as well as machine (statistical) learning.^56^ Because current machine learning (ML) approaches provide correlational but not causal information, they are not explanatory and, consequently, their actual predictability (also known as overfitting) is rather poor.^32, 57^ ML is associated with big datasets, which assume static (stable) relationships and tend to use aggregate (not personalized) data.^58^

According to their transparency and interpretability, machine learning methods are categorized as ‘black’ or ‘white box.’ While black box models are not transparent, they have been reported to be highly informative when complexity is captured; yet, white box models are favored because they can be interpreted.^59^ Both ML versions share two aspects: (i) dependency on large numbers (a possible limitation when *n*=1), and (ii) linearity and reductionism, including the assumption that causes are independent, not interdependent.^50^

The complexity-oriented method differs from the two research traditions mentioned above: while new ‘white box’ approaches have recently been applied in machine learning-related research on antibiotics, they are still influenced by linear models.^57^ In contrast, the non-reductionist approach discriminated regardless of any data distribution because it was based on 3D data shapes (patterns that do not depend on numerical cutoffs). Because biomedicine is complex, dynamic, non-linear, and composed of interdependent processes, the method here described can complement both black- and white-box approaches, countering the overfitting attributed to machine learning.^60^ This approach can also be applied with other dimensions, including molecular and genomic data.

### Clinical applications

#### I. Personalized medicine

The non-reductionist approach circumvents both the risks associated with ambiguity and the limitations of research designs that depend on population metrics, which tend to ignore co-morbidities. Because co-morbidities are associated with poly-pharmacy and exposure to diverse environments and lifestyles, patient-centered (not population-based) approaches are needed, followed by extensive evaluations, i.e., studies on external validity or generalizations.^61, 62^

While co-morbidity (two of more medical conditions diagnosed in the same person) has been reported in 23% of the US general population, and more than 60% of the population above 65 years of age^63^, randomized clinical trials (RCT) assume that only one medical condition exists.^64^ Yet, infectious and non-infectious syndromes –such as tuberculosis, human immunodeficiency virus, and malaria, as well as diabetes mellitus and tuberculosis− tend to yield worse outcomes and affect more people than when they act alone.^65, 66, 67^ Because up to 81.3% of RCT have excluded patients presenting with co-morbidities, the external validity of such designs is unknown. Because they assume that no interactions exist, RCT also lack construct validity.^68, 69^

#### II-Pathogenesis- and vaccine development-related research

As previously reported, the N/L ratio reached high levels in bacterial pneumonia.^70^ A remarkable decrease of this metric was noticed before treatment started (from ∼90 to ∼15 N/L values, Fig. 3G). This was a desirable outcome because, in pneumonia, mortality may follow a protracted inflammation, which may remain even after bacterial clearance.^71^

The observed isoniazid-related immunomodulation of monocytes (Fig. 5 I) may be of interest in vaccine development against tuberculosis. At least two factors may explain the poor efficacy of vaccines against tuberculosis^31^: (i) evaluations based on aggregate data, which promote confounding because they cannot distinguish poor immunogens from poor responders; and (ii) evaluations that measure isolated structures (e.g., a specific cell type) but do not assess functions. In contrast, this study utilized personalized data to investigate multicellular interactions. Thus, the research design here described may investigate individual-level factors which, in tuberculosis, may influence vaccine efficacy.^72^

Antibiotic-leukocyte temporal interactions differed among leukocytes: a double linear process (first, decreasing; later, increasing) was observed between the M% and time but the early linear relationship was not observed when either the L% or N% were investigated over time (Figs. 5 E-G). Hence, the evidence indicated modulation of monocytes: the M% rapidly and linearly decreased in blood before isoniazid-related treatment was prescribed and only increased, also linearly, after that therapy was applied.

The findings reported in Fig. 5 also corroborated a study that related blood monocytes to *ex vivo* protection against tuberculosis. The same report attributed an informative role to the L/M ratio.^73^ Because the L/M ratio distinguished data subsets across five patients and syndromes (Figs. 1-5), its external validity was documented.^50^

#### III - Antibiotic- and antibiogram-related applications

Classic antibiograms (or antibiotic sensitivity tests) have six limitations. They include lack of (i) *in vivo*, (ii) dynamic, (iii) personalized, and (iv) immunological information, and also (v) delayed results and (vi) procedures that do not facilitate the real-time evaluation of therapies.^36^

Furthermore, *in vitro* and *in vivo* systems are likely to differ, e.g., the concentration of iron is rather high *in vitro* but low *in vivo*.^74, 75^ Because antibiograms ignore *dynamics* and depend on growth inhibition, they cannot investigate antibiotic bactericidal activity in drug-tolerant bacteria.^76,77^ Because these tests do not inform at personalized level, they cannot explore immunomodulation. Yet, the 7D approach may help to understand antimicrobial resistance. For instance, amoxicillin-resistant pneumococci and penicillin-resistant *S. aureus* may escape phagocytosis due to direct and indirect actions of these bacteria on neutrophils.^78, 79^ It has been claimed that the first step toward new strategies against antimicrobial resistance is the development of new antibiograms, which should be personalized.^80^ Furthermore, the *long turnaround* (48-72 hours) of antibiograms promotes empirical antibiotic therapy (which may ineffective) and fosters antimicrobial resistance.^80, 81^

Findings support the view that, when ambiguity is prevented, the six problems described above could be avoided. In addition, antimicrobial agent-associated immunomodulation could be investigated. The fact that amphotericin modulated lymphocytes in a co-morbidity case (Fig. 6 G) corroborated (with *in vivo* data) *in vitro* reports on the effect of this antibiotic on immunosuppressed patients affected by fungal infections.^82, 83^ While *in vivo* synergism between amphotericin and linezolid has been reported in studies conducted with flies, to the best of our knowledge, this is the first study that reports *in vivo*, human-based evidence of synergic interactions involving amphothericin, linezolid, and flucytosine.^84, 85^ Because numerous interactions may occur when antibiotic, antiviral, antifungal and/or antituberculosis drugs are prescribed, *in vivo* and personalized monitoring of therapies are needed.^86, 87^

In conclusion, findings supported the construct, internal, and external validity of a highly multi-dimensional method that, *in vivo*, prevents ambiguity, provides patient-specific (personalized) and immunologically explicit (white box) information on infection disease-related dynamics of antibiotic-mediated immuno-modulation. To determine its replicability, additional studies are recommended.

## Data Availability

Data can be shared upon request

## Author contributions

Conceived the study: AR, MR. Contributed data: MI and JF. Wrote the paper: AR, PK, and RD.

