## Supplementary material for "Personalized and dynamic antibiograms-an exploration in seven infectious syndromes": Suppl .Table and Figures

**This PDF file includes:**

Table S1

**Table S1. Presumptive diagnoses, leukocyte profiles and anti-microbial treatments. <**

| Patient ID | Day | Dx | N% | M% | L% | Antimicrobials* |
|---|---|---|---|---|---|---|
| 3 | 0 | SSTI | 82 | 7 | 11 | * |
| 3 | 1 | SSTI | 75.5 | 8.1 | 16.3 | * |
| 3 | 2 | SSTI | 62.9 | 8.6 | 28.3 | * |
| 3 | 3 | SSTI | 48.9 | 14.1 | 36. | * |
| 3 | 4 | SSTI | 53.3 | 12.2 | 34.4 | CRO, VAN |
| 3 | 13 | SSTI | 48.9 | 7.1 | 43.8 | CRO, VAN |
| 10 | 1 | IAI | 74.4 | 10.2 | 15.3 | CIP, metronidazole |
| 10 | 14 | IAI | 69.6 | 8.1 | 22.2 | CRO, metronidazole |
| 10 | 15 | IAI | 87.9 | 4.3 | 7.6 | CRO, metronidazole |
| 10 | 19 | IAI | 76.5 | 10.2 | 13.2 | CRO, metronidazole |
| 10 | 20 | IAI | 78.5 | 8.1 | 13.2 | DOX, CRO, metronidazole |
| 10 | 21 | IAI | 79.7 | 9.1 | 11.1 | DOX, CRO, metronidazole |
| 10 | 26 | IAI | 83 | 7 | 10 | DOX, CRO, metronidazole |
| 10 | 52 | IAI | 78.7 | 6.1 | 15.1 | CRO, metronidazole |
| 18 | 0 | UTI/mening | 80.4 | 9.2 | 10.3 | CRO, ACV: |
| 18 | 1 | UTI/mening | 83.8 | 8.1 | 8.1 | CRO, ACV, AMB |
| 18 | 2 | UTI/mening | 81.8 | 13.1 | 5.1 | CRO, ACV, AMB |
| 18 | 3 | UTI/mening | 82 | 15 | 3 | AMB, SXT, FLC |
| 18 | 4 | UTI/mening | 86.1 | 8.9 | 4.9 | AMB |
| 18 | 5 | UTI/mening | 81 | 12 | 7 | AMB |
| 18 | 6 | UTI/mening | 80.8 | 9.5 | 9.5 | AMB, 5FC, CRO |
| 18 | 7 | UTI/mening | 85.2 | 4.2 | 10.5 | FEP, LZD, AMB, 5FC |
| 19 | 0 | Endoc | 93 | 4 | 3 | VAN, IMP, TZP |
| 19 | 1 | Endoc | 83 | 11 | 6 | VAN, CFZ |
| 19 | 2 | Endoc | 82 | 12 | 6 | VAN |
| 19 | 3 | Endoc | 73.7 | 15.1 | 11.1 | * |
| 19 | 15 | Endoc | 63.2 | 14.2 | 22.4 | * |
| 19 | 16 | Endoc | 68.3 | 13.2 | 18.3 | * |
| 19 | 17 | Endoc | 70.4 | 14.2 | 15.3 | * |
| 19 | 18 | Endoc | 72.7 | 12.1 | 15.1 | * |
| 19 | 19 | Endoc | 73.4 | 12.2 | 14.2 | * |
| 19 | 20 | Endoc | 72.1 | 13.4 | 14.4 | * |
| 19 | 21 | Endoc | 72.1 | 12.3 | 15.4 | * |
| 19 | 26 | Endoc | 74.4 | 13.2 | 12.2 | CFZ |
| 21 | 0 | Pneum | 84.3 | 14.5 | 1.0 | * |
| 21 | 9 | Pneum | 83.1 | 11.8 | 4.9 | * |
| 21 | 11 | Pneum | 88 | 7 | 5 | VAN, TZP, OTV, AZM |
| 21 | 18 | Pneum | 35.0 | 9.2 | 55.6 | MEM, GCV, MFG |
| 21 | 25 | Pneum | 57.3 | 5.6 | 37.0 | MEM, GCV,MFG |
| 21 | 35 | Pneum | 69 | 7 | 24 | MEM, GCV,MFG |

>Table S1. (cont'd) <

| Patient ID | Day | Diagnosis | N % | M % | L% | Antimicrobials |
|---|---|---|---|---|---|---|
| 21 | 37 | Pneum | 64 | 10 | 26 | MEM, GCV,MFG |
| 21 | 38 | Pneum | 74.7 | 8.0 | 17.1 | GCV, TZP, AZM |
| 30 | 0 | Syph | 61.6 | 9.0 | 29.2 | VAN, MXF[#] |
| 30 | 1 | Syph | 62.8 | 9.2 | 27.8 | * |
| 30 | 2 | Syph | 48.9 | 9.3 | 41.6 | ATM, VAN |
| 30 | 3 | Syph | 59.1 | 7.5 | 33.3 | * |
| 30 | 4 | Syph | 51.0 | 9.5 | 39.3 | CLI, ATM, SXT |
| 30 | 16 | Syph | 40.6 | 8.3 | 51.0 | * |
| 33 | 0 | Tuberc | 85 | 10 | 5 | TZP, DOX, CRO, VAN |
| 33 | 1 | Tuberc | 87 | 8 | 5 | DOX, CRO |
| 33 | 2 | Tuberc | 87 | 7 | 6 | * |
| 33 | 3 | Tuberc | 93 | 5 | 2 | * |
| 33 | 4 | Tuberc | 91 | 5 | 4 | RIF, INH, PZA, EMB |
| 33 | 7 | Tuberc | 90 | 6 | 4 | * |
| 33 | 13 | Tuberc | 84.2 | 8.4 | 7.3 | RIF, INH, PZA, EMB |
| 33 | 13 | Tuberc | 75.2 | 12.9 | 11.8 | * |
| 33 | 22 | Tuberc | 75.5 | 14.2 | 10.2 | * |
| 34 | 0 | Endoc | 87.8 | 4.0 | 8.0 | NEI--NAP |
| 34 | 1 | Endoc | 85 | 5 | 10 | NEI--NAP |
| 34 | 4 | Endoc | 84.8 | 4.0 | 11.1 | NEI--NAP |
| 34 | 5 | Endoc | 81.8 | 5.0 | 13.1 | NEI--NAP |
| 34 | 6 | Endoc | 78.7 | 6.0 | 15.1 | NEI--NAP |
| 34 | 7 | Endoc | 69.6 | 6.0 | 24.2 | NEI--NAP |
| 34 | 15 | Endoc | 73.4 | 8.1 | 18.3 | NEI--NAP |
| 34 | 36 | Endoc | 75.7 | 8.0 | 16.1 | NEI--NAP |
| 45 | 0 | SSTI | 90 | 6 | 4 | NAF |
| 45 | 1 | SSTI | 84.8 | 7.0 | 8.0 | NAF |
| 45 | 2 | SSTI | 84 | 8 | 8 | NAF |
| 45 | 3 | SSTI | 86 | 6 | 8 | NAF |
| 45 | 4 | SSTI | 82.4 | 8.3 | 9.2 | NAF |

SSTI: skin/soft tissue infection; IAI: intra-abdominal infection; UTI/mening: urinary tract infection/meningitis; Endoc: endocarditis; Pneum: pneumonia; Syph: syphilis; Tuberc: tuberculosis. N: neutrophil; M: monocyte; L: lymphocyte. ACV: acyclovir; AMB: amphotericin B; ATM: aztreonam , AZM: azithromycin; CFZ: cefazolin; CLI: clindamycin; CIP: ciprofloxacin; CRO: ceftriaxone ('Rocephin');_DOX: doxycycline; EMB: ethambutol; FEP: cefepime; FLC: fluconazole; GCV: ganciclovir; IPM: imipenem; INH: isoniazid; LZD: linezolid; MEM: meropenem; metronidazole, MFG: micafungin; MXF: moxifloxacin; NAF: nafcillin; OTV: oseltamivir ('Tamiflu'); PZA: pyrazinamide; RIF: rifampin; SXT: trimethoprim-sulfamethoxazole ('Bactrim'); TZP: piperacillin-tazobactam ('Zosyn');VAN: vancomycin; 5FC: flucytosine. NEI—NAP: No evidence of infection, no antimicrobial prescribed. [#]: empirical treatment awaiting definitive diagnosis.

>Supplementary Fig. 1<

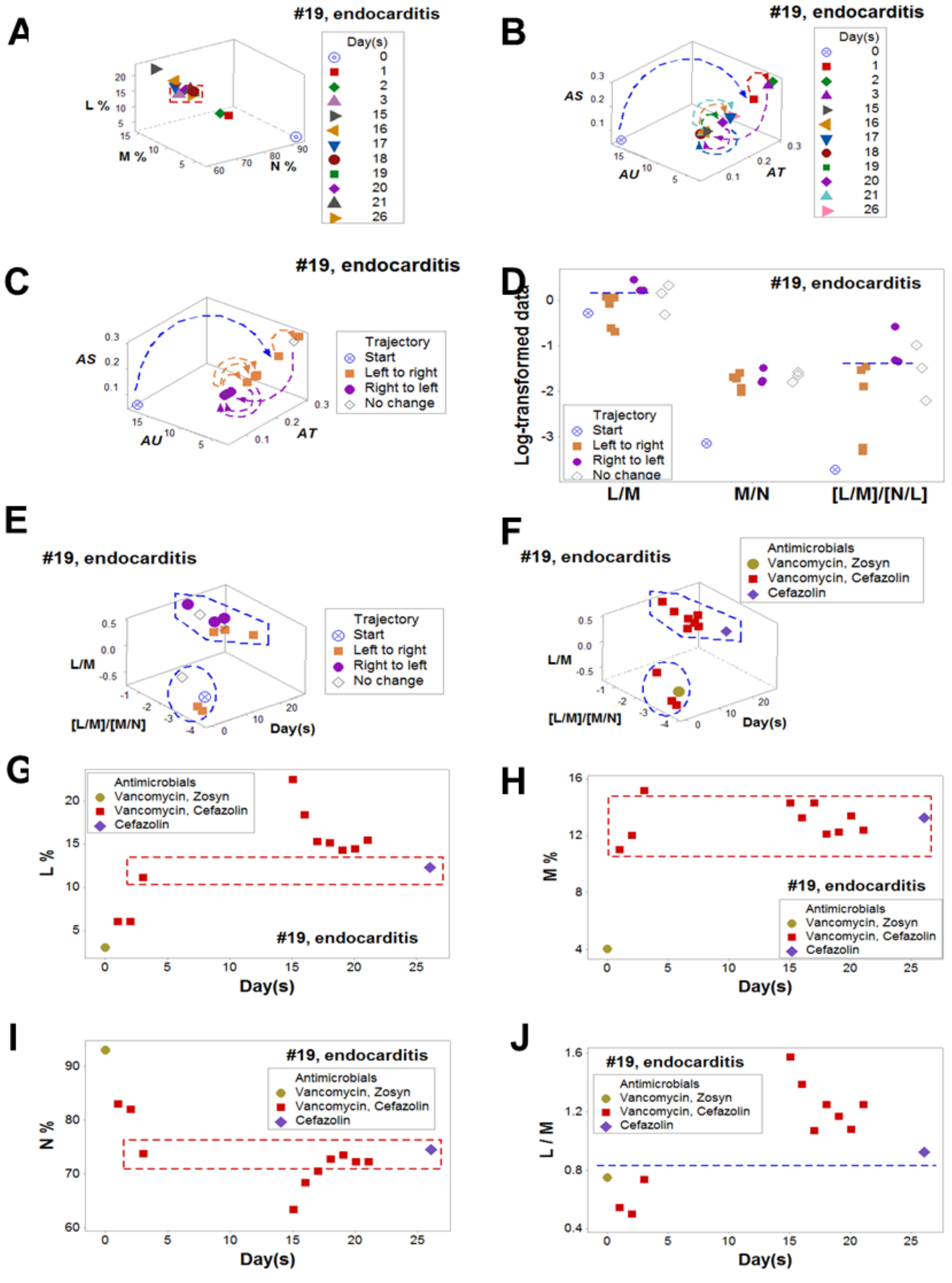

**<Supplementary Fig. 2>**

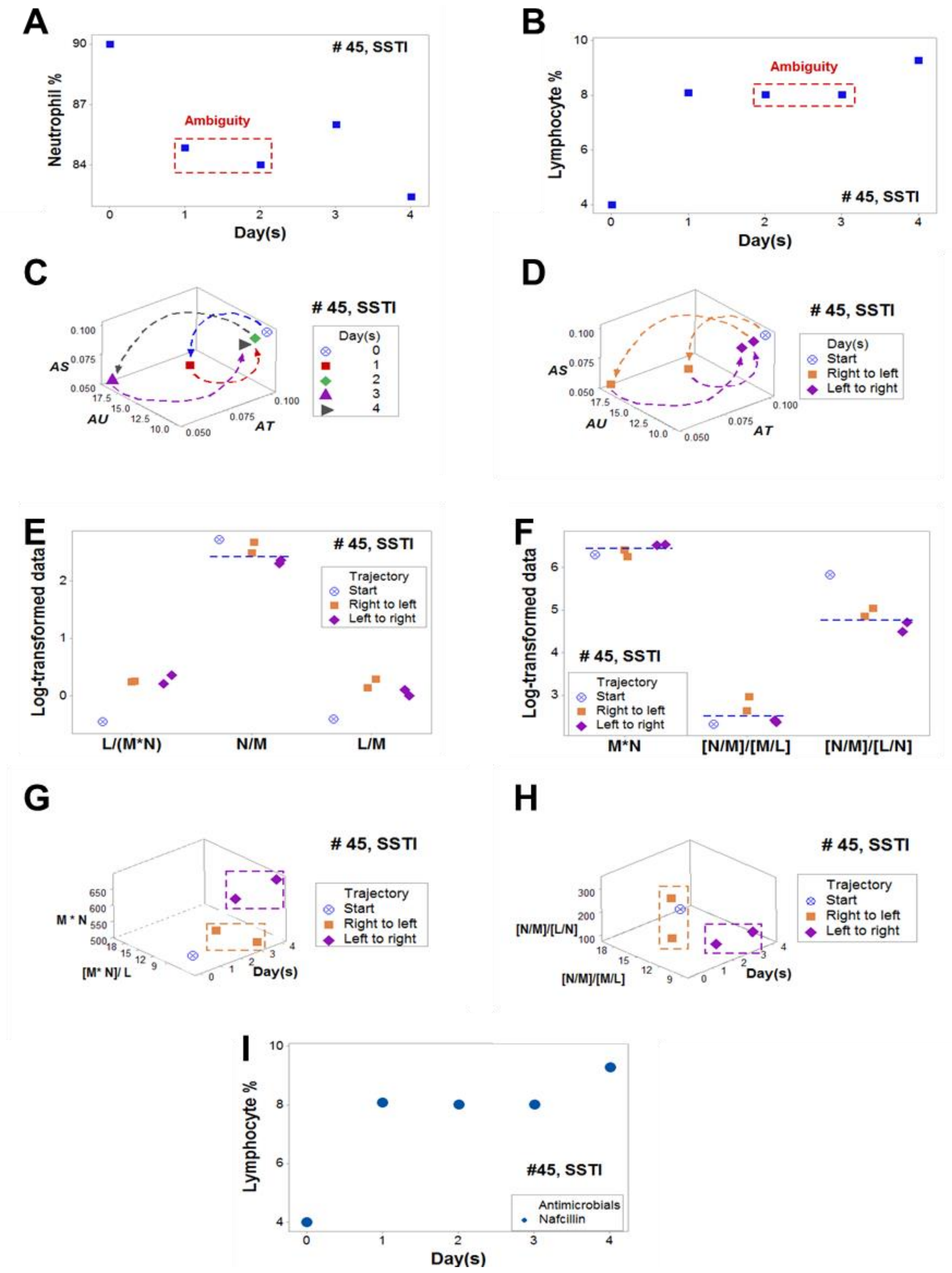

**<Supplementary Fig. 3>**

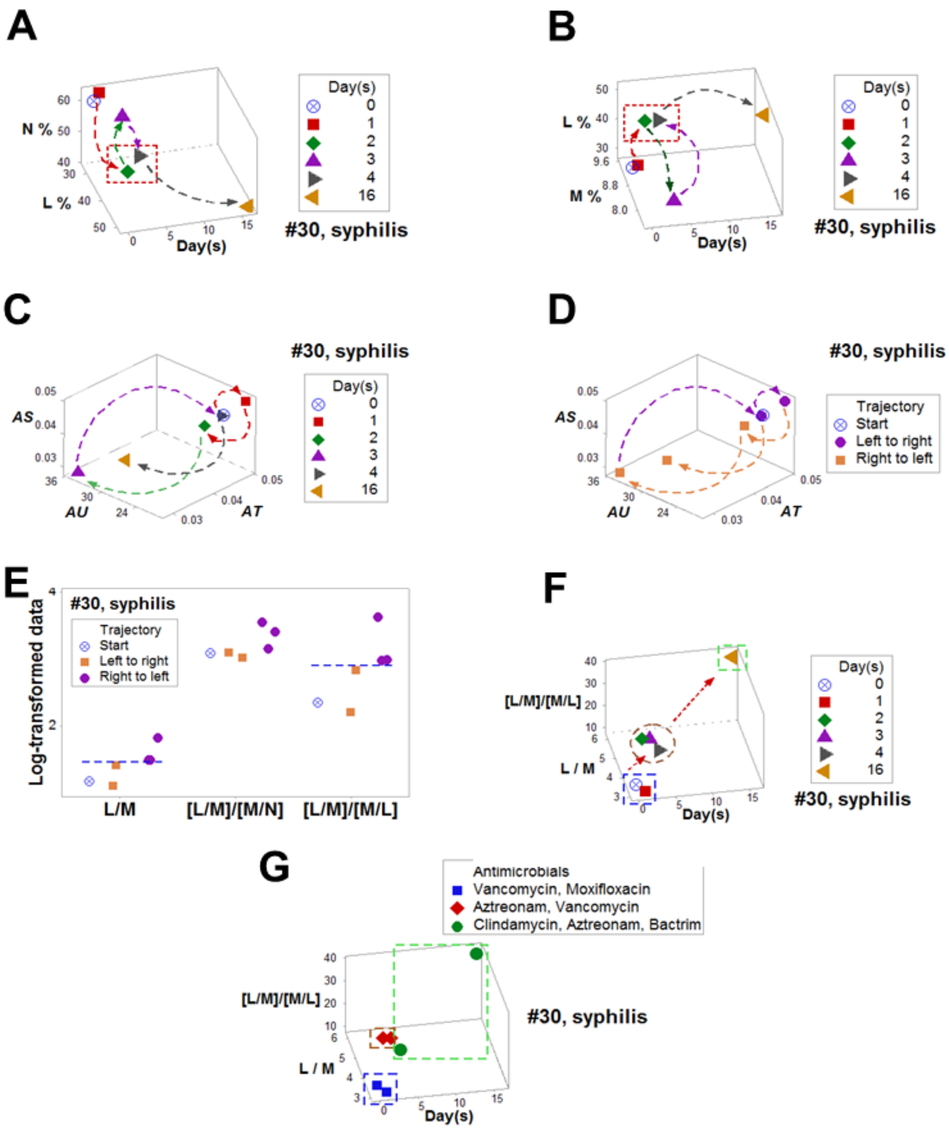
